# Dental Caries Prevalence and Prevention Behaviour Related Factors among Adolescents in Nigeria: A Systematic Review and Meta-analysis

**DOI:** 10.1101/2024.04.18.24305648

**Authors:** Adebukunola O. Afolabi, Adebola O. Ehizele, Ukachi Chiwendu Nnawuihe, Francisca O. Nwaokorie, Ucheoma Nwaozuru, Foluso Owotade, Joanne Lusher, George Uchenna Eleje, Omolola T. Alade, Folahanmi T. Akinsolu, Oliver C. Ezechi, Morenike O. Folayan

## Abstract

**Background:** Adolescence is a critical period of life that has implications for oral health and wellbeing, however, the oral health profile of adolescents in Nigeria is undocumented. This systematic review and meta-analysis determined the prevalence of caries and identified the caries prevention behaviours among adolescents in Nigeria.

**Methods:** In August 2023, a literature search was conducted across PubMed, Google Scholar, Academic Info, Cochrane and Refseek databases to identify studies on dental caries prevalence and caries prevention behaviour among adolescents aged 10 to 19 years in Nigeria. Two reviewers used an independent and double-blind approach to extract data, with discrepancies resolved by a third reviewer. Heterogeneity was assessed using I^2^ percentages, and a funnel plot was constructed to assess the potential for publication bias. The meta-analysis employed a random effects model to ascertain the prevalence of dental caries. Subgroup analysis was conducted to explore the variation in caries prevalence by sex. The review process followed the Preferred Reporting Items for Systematic Reviews and Meta-Analyses (PRISMA) framework guidelines while the Joanna Briggs Institute Critical Appraisal Checklist was used to critically appraise the quality and integrity of the included studies. Registration of this systematic review was completed on PROSPERO CRD42024458849.

**Results:** Sixteen studies met the eligibility criteria. The pooled caries prevalence among adolescents in Nigeria was 23.0% (95% CI: 16, 30) with a pooled prevalence of 16.0% (p < 0.00001, 95% CI: 9 - 23) for males, and a pooled prevalence of 22.0% (p < 0.00001, 95% CI= 11 - 32) for females. The commonest caries prevention practices reported were daily tooth brushing, use of fluoride containing toothpastes and dental service utilization. Factors associated with caries among adolescents were a history of dental service utilization, poor oral hygiene practices and consumption of refined carbohydrates in-between-meals.

**Conclusion:** The high prevalence of dental caries among adolescents in Nigeria warrants programmatic attention with an emphasis on improving oral hygiene practices and controlling the consumption of refined carbohydrates in-between-meals. Dental service utilization for caries prevention needs to be encouraged.

## Background

Dental caries is a significant global public health problem and ranks among the most prevalent oral health challenges faced by children and adolescents [1]. The global prevalence of dental caries in the permanent dentition is 34.1% for adolescents aged 12-19 years [2]. Adolescents are particularly vulnerable to dental caries [7–11] resulting from several factors including poor adherence to caries prevention measures such as regular dental check-ups and good oral hygiene practices [12]. Optimal adherence to dental caries preventive measures are hampered by inadequate awareness, social and cultural influences, and limited access to oral healthcare services [13]. In addition, gender issues has been observed to shape oral health behaviours among adolescents with females reporting more positive preventive oral health behaviors and physical appearances in comparison to male adolescents. [14, 15]. The gender difference may be due to societal expectations that place emphasis on personal hygiene and appearance for girls [6, 14]. Conversely, boys may prioritize other physical health concerns over oral hygiene, resulting in a lower uptake of oral preventive behaviours [14].

Despite the critical importance of understanding gender disparities in caries prevention behaviours among adolescents, comprehensive data on this subject are scarce, particularly within the Nigerian context. Although multiple studies had been conducted on oral health among different populations of adolescents in Nigeria. The absence of information on the epidemiological profile of dental caries among adolescents in Nigeria makes policy formulation and programme development to address disparities difficult. Yet, adolescents are a critical demographic with unique oral health needs. This gap could exacerbate inequities in accessing and utilizing oral health services, undermining the effectiveness of evidence-based preventive oral health interventions for adolescents. A comprehensive analysis of the specific gender dynamics related to caries prevention behaviours among Nigerian adolescents might provide insights that can help address gender-specific barriers to promoting preventive oral health behaviours. This systematic review and meta-analysis is an attempt to bridge the gap in knowledge, determined caries prevalence, caries prevention behaviours and related factors among adolescents in Nigeria.

## Methods

### Protocol and registration

The protocol of this systematic review and meta-analysis was registered a prior with the International Prospective Register of Systematic Reviews (PROSPERO) with CRD42024458849. The study was reported following the Preferred Reporting Item for Systematic Reviews and Meta-analyses (PRISMA) statement and checklist [16]. Each review stage was performed by two authors (reviewers) using a blinded approach while disagreements were resolved through discussion with a third author.

### Eligibility criteria

All published studies, including grey literature, conducted in Nigeria between January 2013 and December 2022, and reporting prevalence of dental caries and caries preventive behaviour among adolescents aged 10-19 years, were included. Study designs considered eligible for inclusion were cross-sectional, cohort, and case-control studies. Studies were included if they presented available data for at least one of the primary outcomes. Studies were excluded if they did not provide information on the sample size, had inaccurate or unavailable outcome data, featured duplicate samples and review articles. Studies with overlapping data from other included studies, as well as case reports, case series, editorials or reviews devoid of primary data were also excluded. In addition, where only an abstract was available, and no further data were available by contacting authors, studies were excluded.

### Search keywords and strategy

A comprehensive systematic literature search was conducted in five databases: Google Scholar, Academic Info, the Cochrane Library, Refseek, PubMed and Medline using the keywords namely: adolescents; caries prevalence; caries prevention behaviours and Nigeria. The initial search syntax was developed for PubMed and later adapted to fulfill the unique search criteria of the other databases. The reference lists of included studies were hand-searched for any additional papers. Where appropriate, attempts were made to contact authors for further information with a single reminder sent where the initial request unanswered. Searches were done with no language restrictions.

### Study Selection

Studies were further screened for eligibility using the PICOTS framework (Population, Intervention, Comparators, Outcomes, Time, Studies). Table1 highlights the PICOTS framework used for this study [17].

**Table 1:** Eligibility Criteria using PICOTS (Population, Intervention, Comparisons, Outcomes, Time, Studies) framework.

|  |  |
| --- | --- |
| Population | Adolescents age 10-19 resident in Nigeria of all sexes. |
| Intervention/Exposure | Caries prevention behaviour |
| Comparators | Individuals who practice caries preventive behaviour or those who practice caries preventive behaviour more frequently than the intervention group. |
| Outcomes | Prevalence of dental caries |
| Time | 01/01/2013 to 31/12/2022 |
| Studies | Observational studies |

Studies were downloaded to the reference management software EndNote 7.8 and subsequently imported in Rayyan software where duplicate studies were removed. Two authors (A.O.A. and A.O.E.) independently assessed the titles and abstracts of studies that met the inclusion criteria after removing duplicates of search outcomes. Studies that did not meet the inclusion criteria and those that copies of the full text of potentially relevant studies were not accessible were excluded. Two review authors (A.O.A. and A.O.E.) independently assessed the eligibility of the retrieved papers and resolved any disagreements by discussion or recourse to a third review author (U.C.N.).

### Data Extraction

Two independent reviewers (A.O.A. and A.O.E.) independently extracted information related to author name, year of publication, and the publishing journal. Additionally, specific information related to the study design (cross-sectional, cohort, case controls), location and setting of the study were also extracted. Details about the study participants (sample size, age distribution, sex of participants, and other unique characteristics) were extracted. Information about participants’ caries prevention behaviour were also extracted. These details include oral hygiene status, frequency of daily tooth brushing, use of fluoridated toothpaste, frequency of consumption of refined carbohydrate daily and dental service utilization. Data on dental caries prevalence were also extracted while results or study outcomes indicating relationship between the prevalence of dental caries and the caries prevention behaviour were recorded, discrepancies were resolved by a third reviewer (U.C.H.).

### Quality and Risk of Bias Assessment

The Joanna Briggs Institute (JBI) Critical Appraisal Checklist [18] was used to appraise the quality of included studies. Each of the eight items of the JBI checklist has options ‘Yes’, ‘No’, ‘Unclear’, ‘Not applicable’. Options ‘Unclear’ or ‘Not applicable’ was scored ‘0’ point, option ‘No’ was scored 1 point while option ‘Yes’ was scored 2 points. The total scores were summed up to a maximum of 16 points and a minimum of ‘0’ point. Any article with total score range 13 to 16 was categorized as ‘high quality’, total score 9 to 12 was categorized as ‘moderate quality’ while total score of 8 and below was categorized as ‘poor quality’. The quality and risk of bias assessment was done by three authors (A.O.A., M.O.F. and F.O.N) while discrepancies were resolved by one of the authors (U.N).

### Primary and Secondary Study Outcomes

The primary outcomes of the review and meta-analysis are the prevalence of dental caries and caries preventive behaviours while the secondary outcomes were factors related to caries prevalence among adolescents in Nigeria.

### Statistical Analysis

Data were analyzed using RevMan 5.4.1 (The Nordic Cochrane Center, Copenhagen, Denmark). First, data was pooled data for the prevalence of dental caries, 95% confidence interval (CI) was used as the effect size and the inverse variance method (Generic Inverse Variance) was selected to calculate the pooled effect. In this statistical procedure, the rate difference (RD) and its standard error are equivalent to the effect of a single rate and the standard error. Cochran’s (Q) statistic test and Higgins and Thompson’s (I^2^) statistic were used to assess heterogeneity between studies. A p-value less than 0.05 was considered statistically significant for the Q-statistics test, and an I^2^ value above 50.0% represented significant heterogeneity. For all other tests, except heterogeneity testing, p-value less than 0.05 were considered statistically significant.

#### Publication Bias

The risk of publication bias was assessed by comparing the characteristics and results of published and unpublished studies included in the current systematic review using the trim-and-fill method to detect asymmetry or missing data in the meta-analysis. This is addition to applying quality appraisal and sensitivity analysis to the included studies in order to assess the methodological quality and risk of bias of each the study included in the review.

#### Sensitivity analysis

The inclusion/exclusion criteria ensure robustness of included articles. Quality assessment of included studies were also undertaken in which studies of lower quality or with a higher risk of bias were excluded. Different statistical models and methods were employed to analyze the data obtained while heterogeneity of reviewed articles were assessed by conducting subgroup analyses to explore potential sources of variability.

## Results

### Selection of studies

As shown in the PRISMA flowchart outlining the results of the literature searches (Figure 1), 1053 records were retrieved. After removing duplicates, 1044 records remained for eligibility screening based on title and abstract. Of these, 1014 studies were excluded based on title and abstract. After the review of the full-text records for the remaining 30 studies, 16 studies met the study inclusion criteria [19–34].

**Figure 1:**
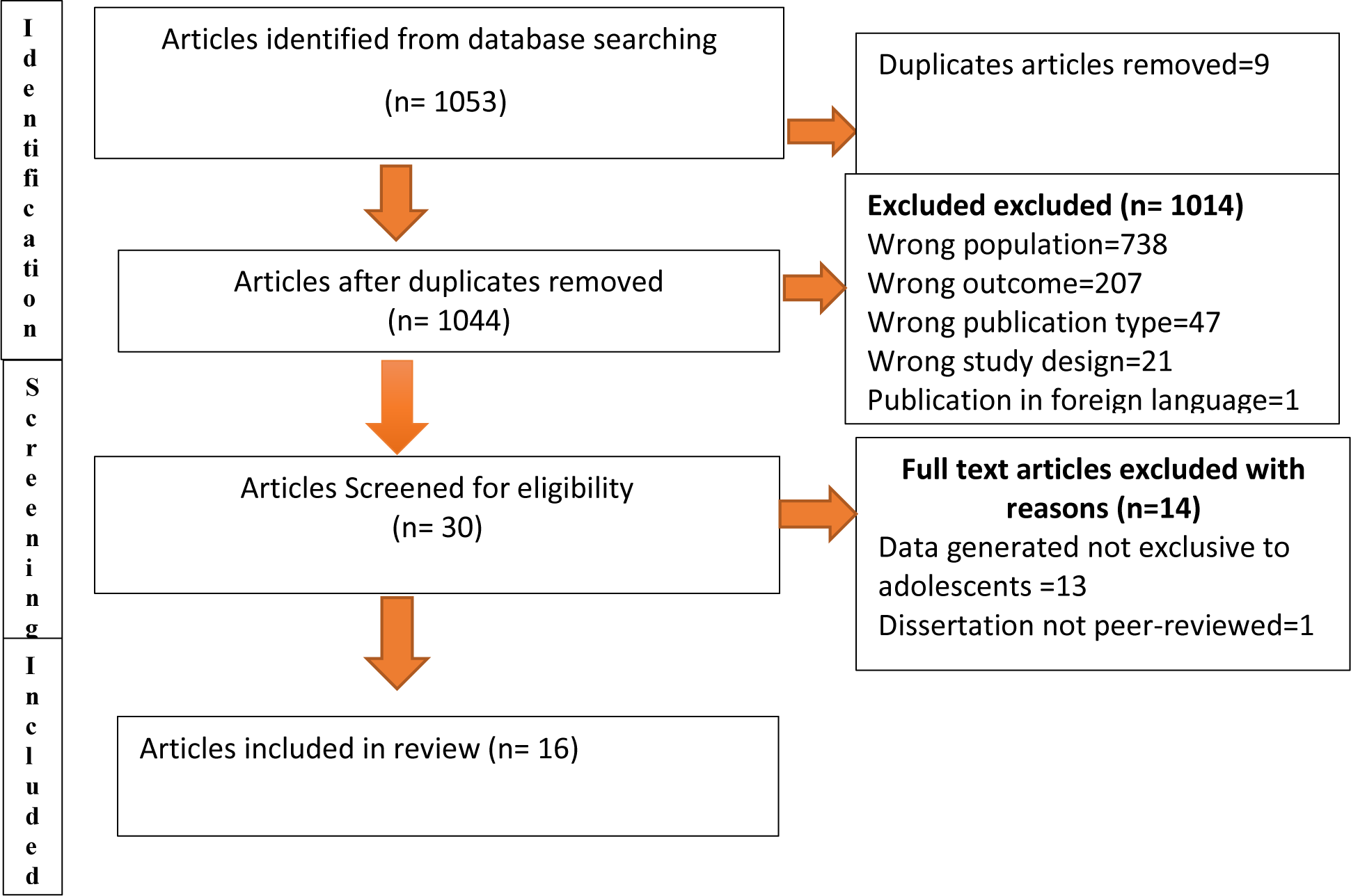
Flow diagram of studies included in the systematic review of caries prevention behavior among adolescents in Nigeria

Table 2 shows the scores on the quality of the studies based on the Joanna Briggs Institute Critical Appraisal Checklist for Prevalence Studies. Nine (56.2%) of the reviewed articles were rated as ‘high quality’ [19,20,22,23,25,26,28,29,32] while seven (43.8%) were of ‘moderate quality [21,24,27,30,31,33,34]. None of the studies conducted an adjusted logistic regression analysis to deal with confounding factors.

**Table 2:**
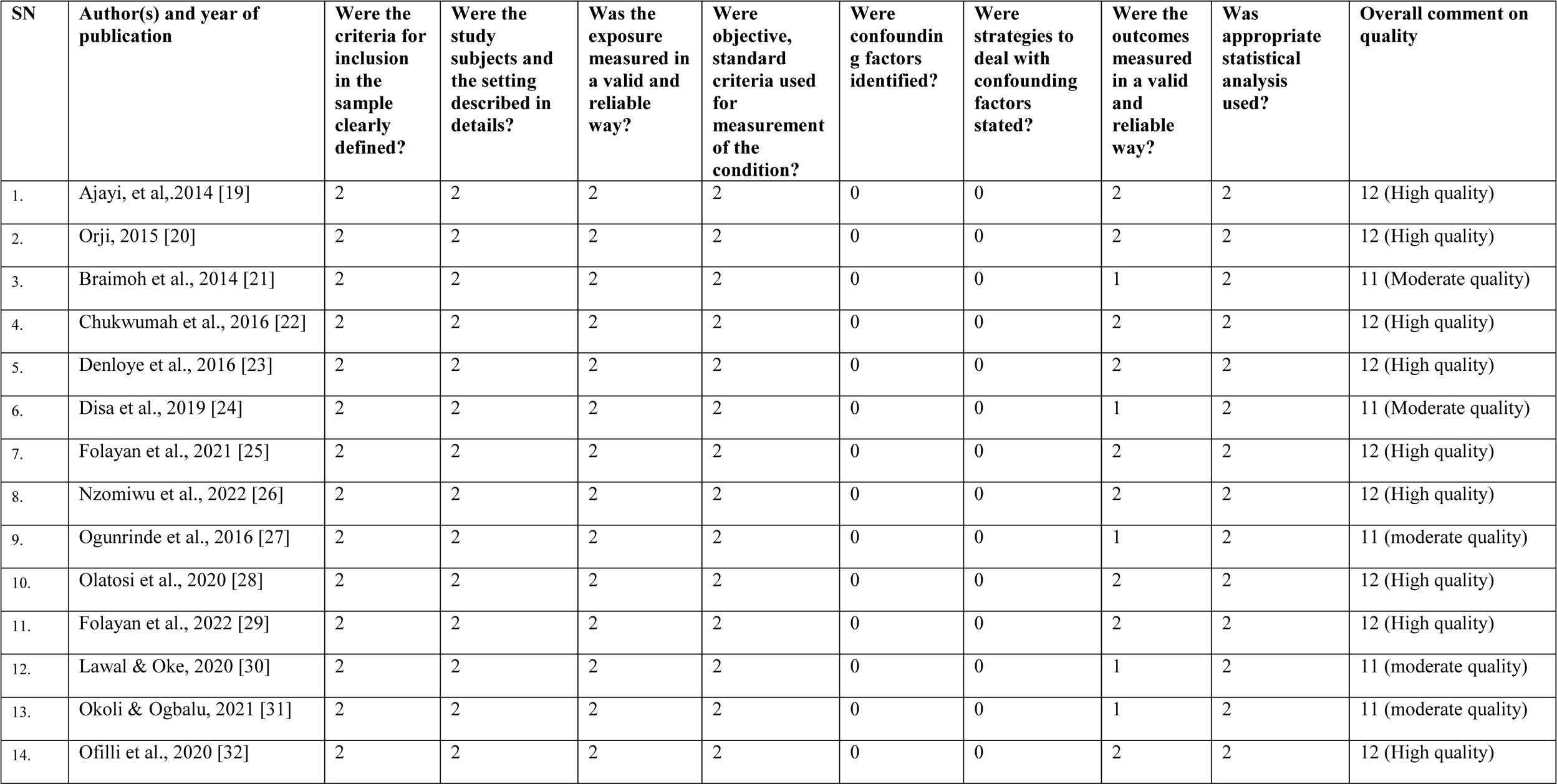
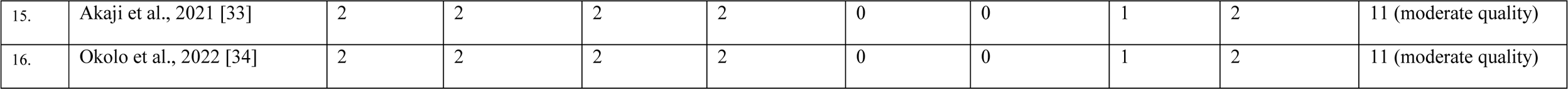
Quality and Risk of Bias Assessment for Reviewed Articles.

### Characteristics of Included Studies

Table 3 shows that all but one of the studies were cross-sectional studies. The exception was a case-control study [24]. The study participants were recruited from primary schools [26], secondary schools [19, 21–23, 27, 28, 30–34], primary and secondary schools [20], hospitals [24] and through household surveys [25, 29].

**Table 3:** Characteristics of Reviewed Articles.

| SN | Authors<br>Year | Study title | Study objective | Study<br>setting | Study location | Sample size | Response<br>rate (%) | Study<br>design | Age of study<br>participants<br>(Years) |
| --- | --- | --- | --- | --- | --- | --- | --- | --- | --- |
| 1. | Ajayi, et<br>al.,2014 [19] | Socio-behavioural risk factors of dental caries among selected adolescents in Ibadan, Nigeria | To determine the influence of sociodemographic and behavioural factors on prevalence of dental caries among adolescents | Secondary schools | Ibadan, Southwest Nigeria | 914<br>Male-451<br>Female-463 | 84.6 | Cross-sectional | 10-19<br>Mean= 14.0± 2SD |
| 2 | Orji, 2015<br>[20] | Prevalence of Dental Caries among Secondary School Children in Ihiala Local Government Area, Anambra State, Nigeria | Determined the prevalence of dental caries among secondary school children | Primary and Secondary schools | Ihiala, Southeast Nigeria | 144<br>Male-64<br>Female-80 | 100.0 | Cross-sectional | 10–19<br>Mean=NR |
| 3 | Braimoh et al., 2014<br>[21] | Caries Distribution, Prevalence, and Treatment Needs among 12–15-Year-Old Secondary School Students in Port Harcourt, Rivers State, Nigeria | To describe caries distribution, prevalence, and treatment needs among school children in Port Harcourt | Secondary Schools | Port Harcourt, Southsouth Nigeria | 195<br>Male-89<br>Female-106 | NR | Cross-sectional | 12–15<br>Mean=13.0±1 SD |
| 4. | Chukwumah et al., 2016<br>[22] | Impact of dental caries and its treatment on the quality of life of 12- to 15-year-old adolescents in Benin, Nigeria | To assess the impact of caries and its treatment on quality of life in 12- to 15-yearold children in Benin, Nigeria | Secondary Schools | Benin, Southsouth Nigeria | 1790<br>Male-949<br>Female-841 | 83.3 | Cross-sectional | 12-15<br>Mean= 14.0 ±1SD |
| 5 | Denloye et al., 2016<br>[23] | Association between dental caries and body mass index in 12-15 year old private school children in Ibadan, Nigeria | To determine dental caries prevalence among a representative sample of 12-15 year old private secondary school students in Nigeria and compare the association of dental caries with body mass index in this sample | Secondary schools | Ibadan, Southwest Nigeria | 595<br>Male -213<br>Female-382 | NR | Cross-sectional | 12-15<br>Mean=NR |
| 6. | Disa et al.,<br>2019 [24] | Predictors of dental caries among adults and adolescents in a dental clinic in north-eastern Nigeria | To determine the predictors of dental caries among adults and adolescents | Hospital-based | Damaturu, Northeast Nigeria | 124<br>Control Case<br>Male-29 30<br>Female-36 29 | NR | Case-control | 12-19<br>Mean=NR |
| 7. | Folayan et al., 2021<br>[25] | Individual and familial factors associated with caries and gingivitis among adolescents resident in a semi-urban community in South-western Nigeria | To describe the prevalence, and individual and familial risk indicators for dental caries and gingivitis among 10–19-year-old adolescents | Households | Ile-Ife, Southwest Nigeria | 1472<br>Male-846<br>Female-626 | NR | Cross-sectional | 10-19<br>Mean=15.0±3SD |
| 8 | Nzomiwu et al., 2022 [26] | Oral Health Knowledge and Behavior among Public Primary Schoolchildren in Lagos, Nigeria | Evaluated the oral health knowledge and behavior among primary school pupils | Primary schools | Lagos, Southwest Nigeria | 434 Male-229 Female-205 | 96.4 | Cross-sectional | 12-19 Mean=12±2SD |
| 9 | Ogunrinde et al., 2016 [27] | Dental care knowledge and practices among secondary school adolescents in Ibadan North Local Government Areas of Oyo State, Nigeria | Assessed the dental care knowledge, and practice of secondary school adolescents | Secondary schools | Ibadan, Southwest Nigeria | 412 Male-249 Female-163 | 100.0 | Cross-sectional | 10-19 Mean=15.0±1SD |
| 10 | Olatosi et al., 2020 [28] | Disparities in Caries Experience and Socio-Behavioural Risk Indicators Among Private School Children in Lagos, Nigeria | Determined the prevalence and socio-behavioural risk factors for dental caries among children at selected LGAs | Secondary schools | Lagos, Southwest Nigeria | 600 Male-307 Female-285 | 98.7 | Cross-sectional | 10–19 Mean=NR |
| 11 | Folayan et al., 2022 [29] | Sexual health risk indicators and their associations with caries status and gingival health of adolescents resident in sub-urban South-west Nigeria | Investigated sexual risk factors associated with caries | Households | Ile-Ife, Southwest Nigeria | 1244 Male-701 Female-543 | 100.0 | Cross-sectional | 10-19 Mean=15.0±3SD |
| 12. | Lawal & Oke, 2020 [30] | Clinical and sociodemographic factors associated with oral health knowledge, attitude, and practices of adolescents in Nigeria | Assessed clinical and sociodemographic factors associated with oral health knowledge, attitude, and practices of adolescents | Secondary schools | Ibadan, Southwest Nigeria | 2097 Male-1126 Female-971 | 99.9 | Cross-Sectional | 12-18 Mean=15.0±1SD |
| 13 | Okoli & Ogbalu, 2021 [31] | Prevention of common oral diseases among Senior Secondary School Students in Enugu State, Nigeria | Ascertained the prevention of common oral diseases among selected Senior Secondary School students | Secondary schools | Enugu, Southeast Nigeria | 900 Male-286 Female-614 | NR | Cross-sectional | 10-19 Mean=NR |
| 14 | Ofilli et al., 2020 [32] | Oral hygiene practices and utilization of oral healthcare services among in-school adolescents in Calabar, Cross River State, Nigeria | Determined the oral hygiene practices, awareness and utilization of oral healthcare services among in-school adolescents | Secondary schools | Calabar, Southsouth Nigeria | 335 Male-189 Female-146 | 95.7 | Cross-sectional | 10-19 Mean=13.0±2SD |
| 15 | Akaji et al., 2021 [33] | Assessing dental caries and related factors in 12-year-old Nigerian school children: report from a Southeastern state | Assess dental caries and related factors using the Decayed Missing and Filled Teeth (DMFT), Significant Caries Index (SiC), Restorative and Met Need Indices in 12-year-old school children in a Southeastern state of Nigeria | Secondary schools | Enugu, Southeast Nigeria | 360<br>Male-226<br>Female-134 | 100.0 | Cross-sectional | 10-19<br>Mean=NR |
| 15 | Okolo et al., 2022 [34] | Dental Caries Prevalence, Severity, and Pattern Among Male Adolescents in Kano, Nigeria | Determined the prevalence, pattern, and severity of caries among 10-12-year-old adolescents | Secondary schools | Kano, Northwest Nigeria | 341<br>Male-341 | NR | Cross-sectional | 10-19<br>Mean=NR |

*NR= Not Reported*

Out of the 16 studies, eight (50.0%) were conducted in the southwest region: four in Ibadan [19, 23, 27, 30], two in Lagos [26, 28], and two in Ile-Ife [25, 29]. In addition, three (25.0%) of the studies were conducted in the Southsouth region of Nigeria: one in Port Harcourt [21], one in Benin City [22] and one in Calabar [32]. Three (18.8%) studies were conducted in the Southeast region: two in Enugu [31, 33] and one in Ihiala [20]. There was a single study (6.3%) conducted in Kano, Northwest Nigeria [34] and in Damaturu, Northeast Nigeria [24] respectively. The study participants recruited were cohorts of adolescents 10-19-years-old [19, 20, 25, 27–29, 31–33], 12-15-years-old [21–23], 12-19-years-old [24, 26] and 12-18-years-old [30].

Eight (50.0%) of the studies [22, 25, 27, 29, 30, 32, 33, 28] conducted a power calculation to justify the sample size, and eight (50.0%) [19, 20, 21, 24, 23, 26, 31, 34] had no evidence of sample power calculation to justify sample size. The participant population size in each study ranged from 124 to 2097 adolescents. The total number of participants recruited in all the studies reviewed was 11,957: 6,325 males and 5632 females. One of the studies exclusively recruited males [34]. In addition, nine of the studies (56.3%) reported on the response rate of participants [19, 20, 22, 26, 27, 28, 29, 30, 32, 33].

#### Prevalence of Caries among Adolescents

Table 4 shows that the prevalence of caries ranged between 3.4% [25] and 54.4% [33]. Ten (62.5%) studies reported prevalence of caries among the adolescents [19–23, 25, 28, 30, 33, 34], four (25.0%) reported gender variations in caries prevalence among the adolescents [19–21, 23], while six (37.5%) neither reported prevalence of caries nor the gender variations among the adolescents [24, 26, 27, 29, 31, 32].

**Table 4:** Caries Prevention Behaviours among Adolescents.

| SN | Authors | Prevalence of Dental Caries |  |  | Caries Prevention Behaviour |  |  |  |  |  |  |
| --- | --- | --- | --- | --- | --- | --- | --- | --- | --- | --- | --- |
|  |  | Total | Male | Female | Other devices | Toothbrush and paste | Tooth cleaning frequency | Use of fluoridated toothpaste | Use of chewing stick | Sugar consumption in drinks/ snacks | Dental service utilization |
| 1. | Ajayi, et al.,201 [19] | 10.6% | 10.4% | 10.8% | Cotton wool and toothpaste: 0.7% | 92.3% | Once daily: 50.2%<br>Twice daily: 44.7%<br>Once in 2 days: 2.4%<br>Weekly: 0.7%<br>No response: 2.0% | - | 8.0% | Sweet drinks: 100%<br>Sugary snacks: In between meals: 95.4%<br>With meals: 2.5% | 25.8% |
| 2 | Orji, 2015 [20] | 45.8% | 34.4% | 55.0% | - | 93.1% | Once daily: 69.4%<br>Twice daily: 22.2%<br>Occasionally: 8.3% | - | 6.9% | Sugary snacks: 73.6%<br>Sweet drink: 72.2% | 33.3% |
| 3 | Braimoh et al., 2014 [21] | 15.4% | 7.2% | 8.2% | - | - | - | - | - | - | 6.3% |
| 4 | Chukwumah et al., 2016 [22] | 21.9% | 6.9% | 15.0% | - | - | - | - | - | - | - |
| 5 | Denloye et al., 2016 [23] | 16.0% | 16.0% | 16.2% | - | - | - | - | - | - | - |
| 6 | Disa et al., 2019 [24] | - | - | - | Other materials:3.2% | 81.5% | <twice daily: 34.7%<br>Twice daily: 65.3% | - | 15.3% | 23.4% | Ever: 59.7% |
| 7 | Folayan et al., 2021 [25] | 3.4% | - | - | Dental floss<br>Daily: 16.1%<br>≥ Not daily:83.9% | - | ≤ once daily: 91.3%<br>≥ twice daily: 8.7% | Always: 91.7%<br>Not always:8.3% | - | Frequent: 63.9%<br>Not frequent: 36.1% | In last 12 months: 1.1% |
| 8 | Nzomiwu et al., 2022 [26] | - | - | - | Mouth wash: 4.4%<br>Cotton wool: 1.6% | 92.2% | Once daily: 36.6%<br>≥twice daily: 63.4% | - | 1.8% | - | Ever: 24.9% |
| 9 | Ogunrinde et al., 2016 [27] | - | - | - | Toothbrush and chewing stick: 3.6%<br>Other material:1.4% | 93.0% | Once daily: 7.8%<br>Twice daily: 89.6% | - | 1.9% | - | 31.6% |
| 10 | Olatosi et al., 2020 [28] | 16.0% | - | - | - | - | Not every day:9.6%<br>Once a day: 47.3%<br>≥twice daily: 35.8%<br>No response: 7.3% | - | - | Sugar snacks: 94.1%<br>Sugar drinks: 89.7% | Ever: 54.2% |
| 11 | Folayan et al., 2022 [29] | - | - | - | - | - | Twice daily: 8.8% | 91.2% | - | Between meals: 62.0% | In last 12 months: 1.1% |
| 12 | Lawal & Oke, 2020 [30] | 4.7% | - | - | - | - | - | 90.2% | - | - | 3.8% |
| 13 | Okoli & Ogbalu, 2021 [31] | - | - | - | Tooth powder: 49.3%<br>Mouthwash: 21.0% | 95.7% | Every morning: 98.9%<br>Every night: 36.1% | 95.7 | 55.9% | 87.8% | 17.6% |
| 14 | Ofilli et al., 2020 [32] | - | - | - | Soap & brush: 0.3% | 98.8% | Once daily: 17.0%<br>≥twice daily: 82.1% | 98.8% | 0.3% | - | 40.6% |
| 15 | Akaji et al., 2021 [33] | 54.4 | -54.9% | -53.7% | - | - | Once daily: 90.0%<br>≥twice daily: 1.16% | - | - | 96.4% | - |
| 16 | Okolo et al., 2022 [34] | 22.9% | - | - | - | - | - | - | - | - | - |

#### Caries Prevention Behaviours Associated with Caries

Table 4 also shows that 13 studies reported caries prevention practices among the adolescents [19–21, 24–33]. Overall, more than 90% of adolescents use toothbrush and toothpaste for tooth cleaning with the highest proportion of 98.8% reported in Calabar, Southsouth Nigeria [32]. Other devices used for tooth cleaning include chewing stick [19, 20, 24, 26, 27, 31, 32], cotton wool [19, 26], dental floss [25], mouth wash [26, 31], soap [32] and other materials [26, 27, 31].

Twelve studies reported on the the frequency of tooth cleaning [19, 20, 24–33]. The proportion of adolescents that brush twice daily or more range from 1.1% [33] to 89.6% [27]. The majority of participants clean their mouth twice daily or more in Damaturu [24], Lagos [26], Ibadan [27], and Calabar [32]. The majority of participants clean their mouth once daily in Ibadan [19], Ihiala [20], Enugu [31, 33], Ile-Ife [25], Lagos [28], and Ibadan [30]. In addition, 36.1% brush at night [31].

The use of fluoridated toothpastes was reported by seven studies [19, 24, 25, 29–32]. More than 90% of respondents used fluoridated toothpastes [25, 29–32] except for the single study in Northern Nigeria where the proportion of respondents who used fluoridated toothpaste was 81.5% [24].

The consumption of refined carbohydrate was reported in seven studies [19, 20, 24, 26, 27, 31, 32]. Overall, the proportion of respondents who consumed refined carbohydrate was more than 60% except for the single report from Northern Nigeria where 23.4% of respondents reported consumption of refined carbohydrate [24]. In addition, 73.6% - 97.9% consume refined carbohydrate in the form of snacks [19, 20] and 72.2% - 100% consume this in the form of sweetened drinks [19, 20, 28]. Up to 62% −95.4% of respondents consume refined carbohydrate in-between-meals [19, 29].

Eleven studies reported on the use of dental services [19–21, 24–32]. The proportion of respondents who use the dental service ranged from 1.1% in the last 12 months [25, 29] to 59.7% of ever used [24].

#### Factors Associated with Prevalence of Caries and Caries Preventive Behaviours

Seven studies did not report on caries preventive behaviour [20–23, 27, 31, 34]. Of the nine studies on the association between the prevalence of caries and caries preventive behaviour, four showed an association as highlighted in Table 5. The caries preventive behaviours associated with caries prevalence were history of dental service utilization [19], oral hygiene practices [24, 28, 30] and consumption of refined carbohydrates between meals [29]. Three studies showed associations between a history of dental service utilization and the use of caries preventive behaviours [26, 30, 32].

**Table 5:** Factors Associated with Caries Prevalence and Caries Preventive Behaviour among Adolescents in Nigeria.

| Variables identified | Studies |
| --- | --- |
| **Factors associated with caries prevalence** |  |
| History of dental service utilization | [19] |
| Frequency of tooth cleaning | [24, 28] |
| Consumption of refined carbohydrates in-between meals | [29] |
| **Factor associated with caries preventive behaviour** |  |
| History of dental service utilization | [26, 30, 32] |

#### Pooled prevalence of dental caries among adolescents in Nigeria

In addition, Figure 2 shows that the pooled prevalence of dental caries among adolescents in Nigeria was estimated as 23.0% (95% CI: 16, 30) based on a random-effects model-based meta-analysis conducted on all data points. In addition, the heterogeneity index of I^2^ = 99% (*P* < 0.0001), confirmed substantial heterogeneity among the included studies.

**Figure 2:**
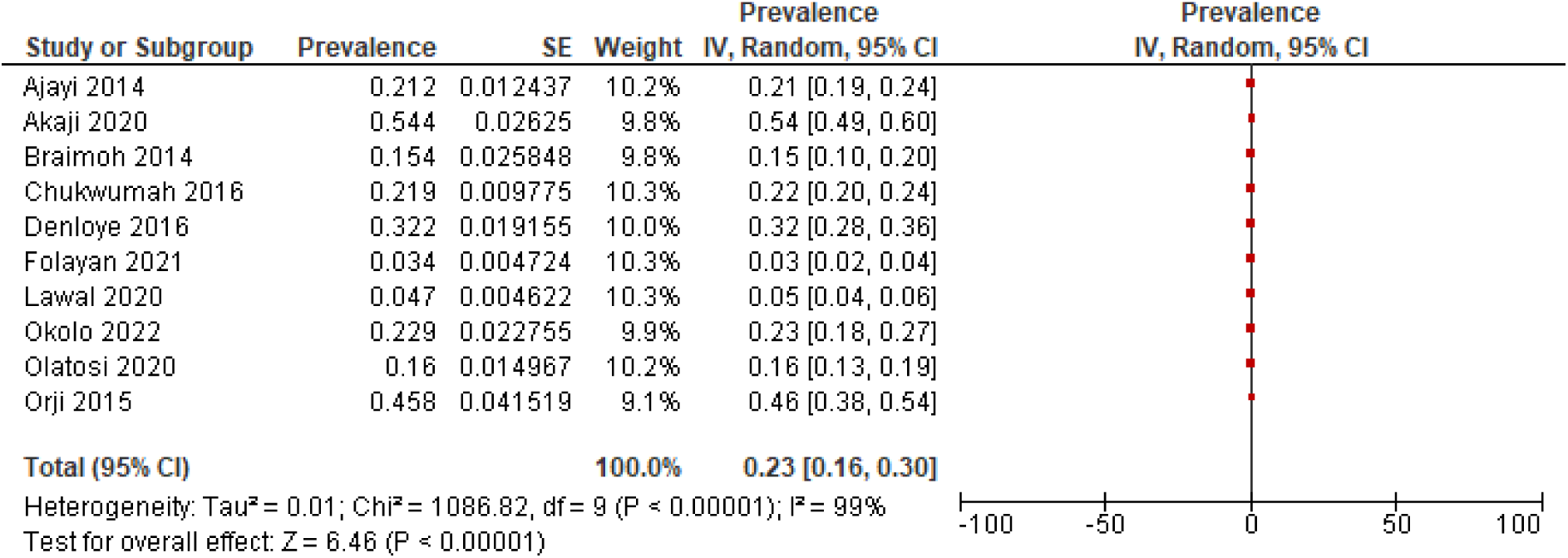
Prevalence of Dental Caries among adolescents in Nigeria

Male participants however had a pooled prevalence of 16.0% (95% CI: 9 - 23) (Figure 3), while female participants had a pooled prevalence of 22.0% (95% CI: 11 - 32) (Figure 4).

**Figure 3:**
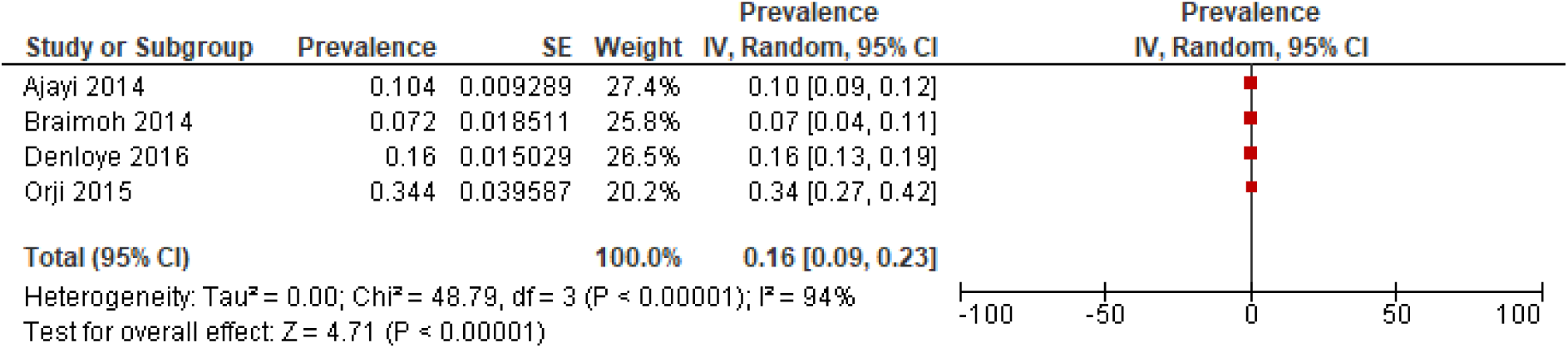
Prevalence of Dental Caries among Male Adolescents in Nigeria

**Figure 4:**
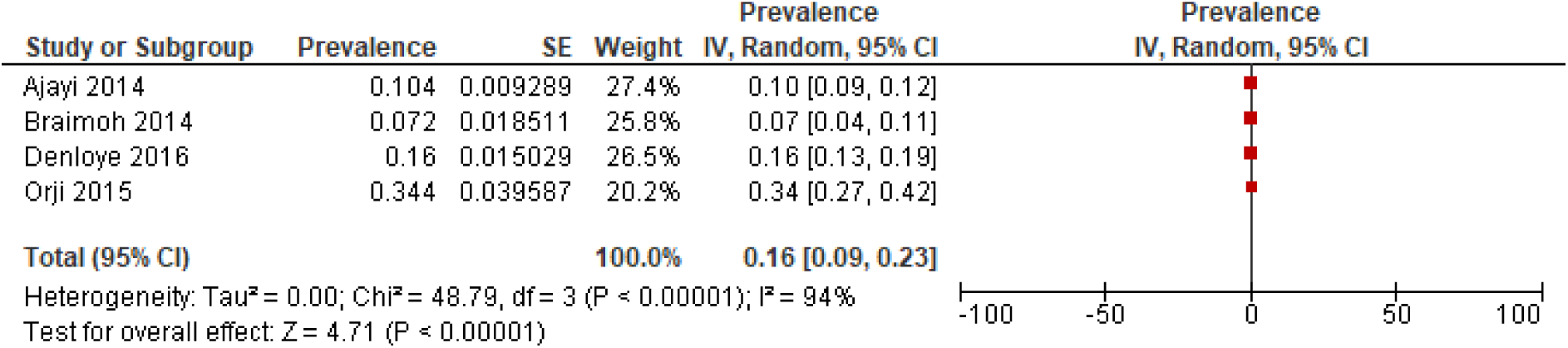
Prevalence of Dental Caries among Female Adolescents in Nigeria

#### Sensitivity Analysis: Impact of Study Quality

Table 2 shows the prevalence estimates and heterogeneity in studies categorized as having high and medium quality. Figure 5 shows the pooled prevalence was found to be 0.23 (95% CI [0.13, 0.34]) for high quality studies while Figure 6 shows that the pooled prevalence was 0.24 (95% CI [0.03, 0.45]) for medium quality. These pooled prevalences were minimally different from the pooled prevalence for all studies was 0.23 (95% CI [0.16, 0.30]) as shown in Figure 2.

**Figure 5:**
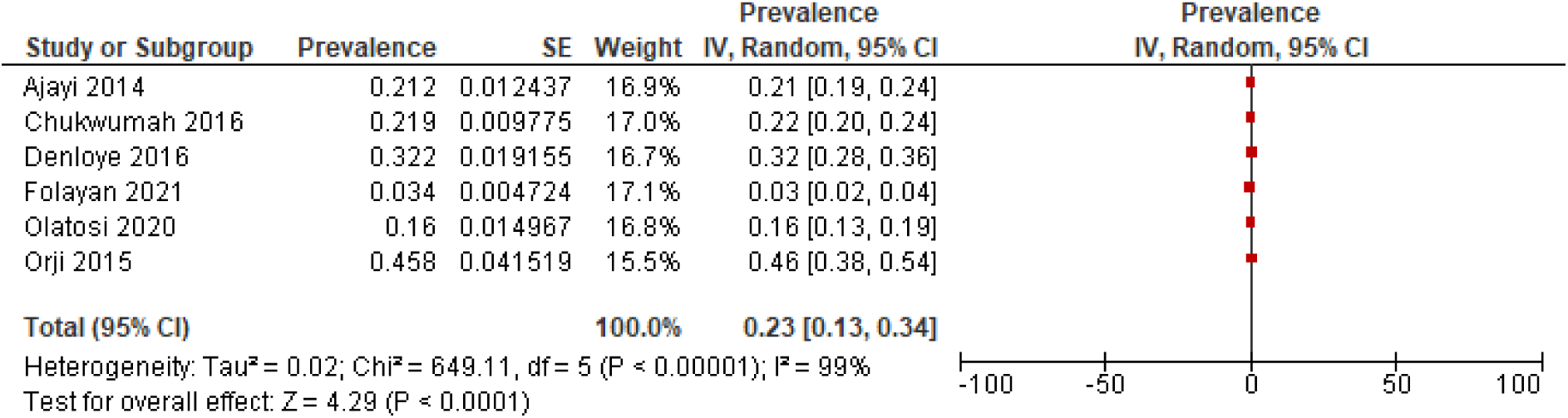
Forest Plot of pooled prevalence of Dental Caries among adolescents in Nigeria using High Quality Assessed Studies

**Figure 6:**
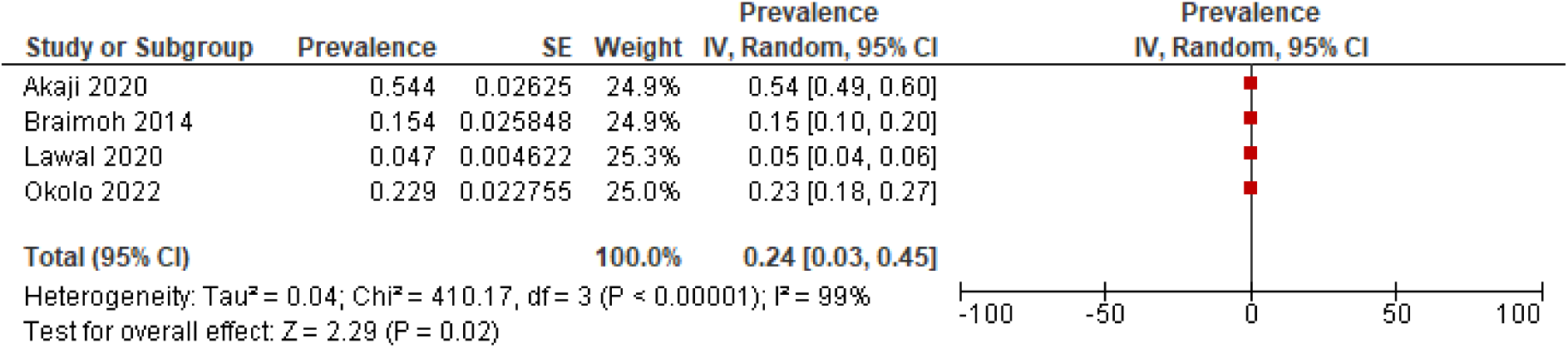
Forest Plot of pooled prevalence of Dental Caries among adolescents in Nigeria using Medium Quality Assessed Studies

#### Publication Bias

Figure 7 shows the symmetric distribution of studies on the funnel plot. This suggests that there is no apparent asymmetry indicative of publication bias. The range of prevalence estimates, along with their corresponding precision measures, is consistent with the expected variability in a well-conducted systematic review.

**Figure 7:**
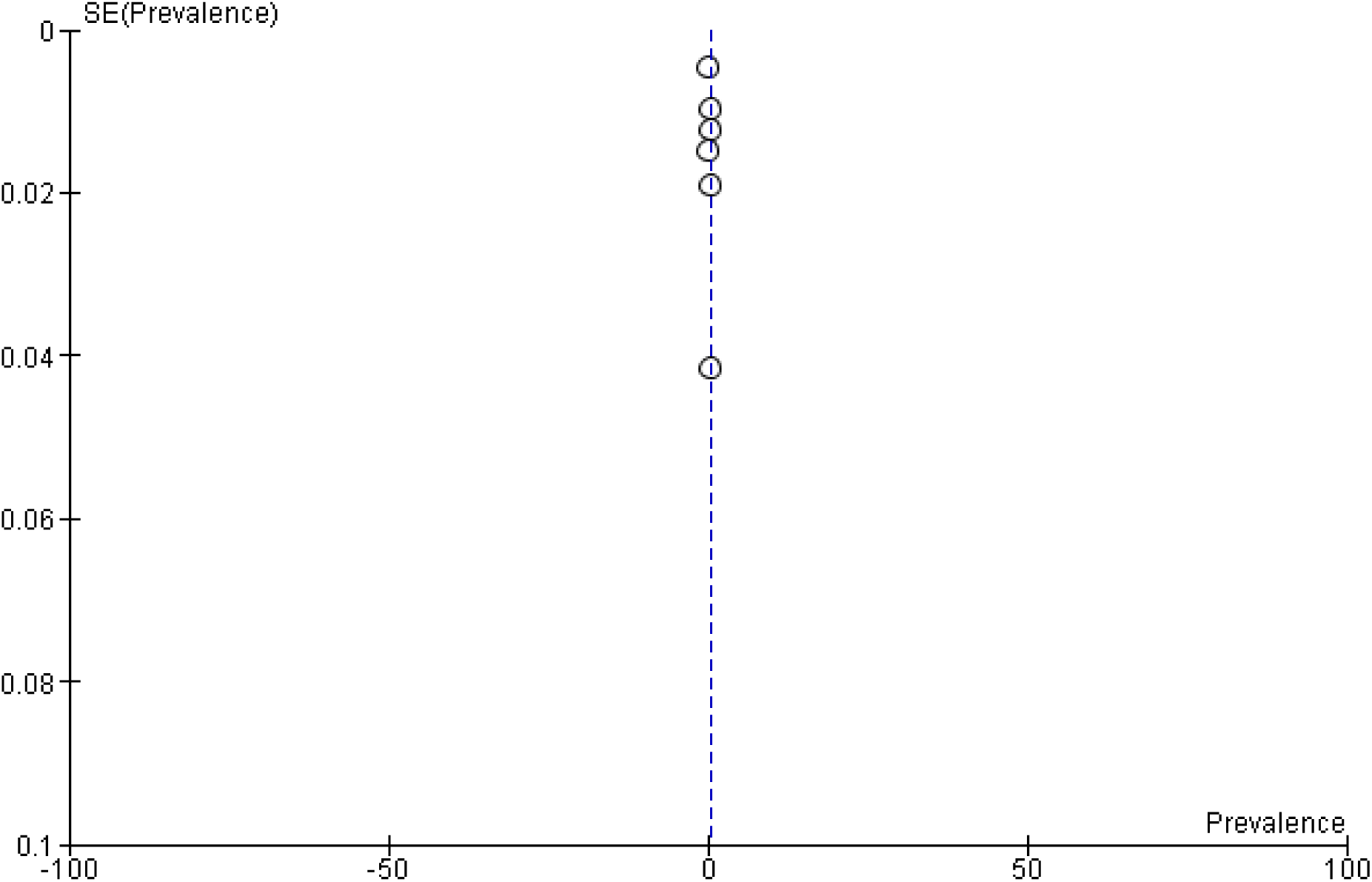
A bias assessment funnel plot of all included studies reporting Dental Caries among adolescents in Nigeria

## Discussion

This systematic review determined the prevalence of dental caries among adolescents in Nigeria, and caries prevention behaviours. The findings suggests that one in four adolescents had dental caries, with a higher risk among females. In addition, studies determining the prevalence of caries among adolescents in Nigeria were mainly conducted in secondary schools, in Southwest Nigeria with sparse information from Northern Nigeria. The study methodology was also a huge concern as half the study had no sample size calculation despite being cross-sectional in design, and none of the studies adjusted for confounders. Although the study had high heterogeneity, and nearly half the study lacked sample power calculation, the findings of the systematic bias analysis suggest that the overall prevalence estimate is robust and not substantially influenced by the exclusion of studies based on their quality assessments. In addition, the symmetric nature of the funnel plot indicates there is no significant publication bias in the meta-analysis. The key prevention behaviours include tooth brushing frequency, dental service history, fluoride toothpaste use, chewing stick, and dental floss. Factors associated with caries include dental service history, tooth brushing frequency, between-meals and consumption of refined carbohydrates.

This first systematic review on adolescent caries in Nigeria was directed by reporting guidelines thereby enhancing the credibility and it sources for peer reviewed articles in five databases and it used the PICOTS framework thereby enhancing inclusivity. Blinded review, the use of RevMan and consideration of heterogeneity, publication bias and sensitivity added rigor and robustness to the study methods. An additional strength of this review was the adherence to a registered protocol developed a priori and the use of a thorough search strategy. Authors were also contacted for additional data where appropriate. Despite these strengths, the study had a few limitations. These include the restricted literature review timeframe (2013-2022); and the potential bias hospital-based data introduced to the study. Despite limitations, the study yielded important findings.

A primary concern lies in the significant heterogeneity in this study. Although the heterogeneity for the meta-analyses were high and statistically significant, the sensitivity analysis and publication bias analysis suggests the observed heterogeneity were not due to methodological diversity or differences in outcome assessments thereby eliminating methodological differences in the pooled studies. The cause of the heterogeneity could, however, not be ascertain in the sensitivity analysis conducted. We, however, recognize concerns with the study design as none of the studies conducted an adjusted regression analysis. This suggests inadequacy in methodological guidance for dental caries research in Nigeria, or shortcomings during the peer review process.

The pooled prevalence of dental caries is high among Nigerian adolescents, especially in females. While this prevalence is lower than the global average of 34.1% for 12-19 year-olds [2], the 66.0% among adolescents in Uganda [35], 65.2% in Eritrea, 57.8% in Sudan and 30.7% in Tanzania [36], this remains a concern. The high caries prevalence indicates broader public health issues such as possible challenges with access to dental care, oral health education, and effectiveness preventive programmes. Comprehensive strategies that include oral health education campaigns in schools, communities, and healthcare facilities that serve adolescents, are needed. Emphasis should be on promoting oral hygiene practices, including regular tooth brushing, and addressing dietary habits, particularly sugary foods and beverages, as highlighted in this study.

An effective measure involves integrating oral health into the existing community health response program. Nigeria has made substantial investments in community health workers for health education and prevention promotion [37]. Advocacy efforts are crucial to incorporate oral health education into these initiatives, emphasizing the importance of twice daily tooth brushing for optimal oral hygiene [38]. This approach aims to reduce the consumption of refined carbohydrates between meals and encourage timely utilization of dental services for preventive care. Currently, dental services are mainly sought for curative purposes, and adolescents lack awareness of preventive care benefits [39–41]. Barriers to healthcare service utilization in Nigeria include poor accessibility, cost, and a cultural reluctance towards preventive care [42]. A comprehensive, multifaceted response is essential to curb caries prevalence in a country where the oral health system is weak and unresponsive to preventive care.

The current study reveals that a significant proportion of adolescents brush their teeth only once a day, whereas evidence suggests that twice daily tooth brushing with fluoridated toothpaste is essential for optimal caries control [43]. Although there is a high proportion of adolescents using fluoridated toothpaste in the study, there is a potential risk of an increase in the use of less expensive non-fluoridated toothpaste. This risk is influenced by public campaigns against fluoride use [44] and the lower costs of non-fluoridated toothpaste [45].

Considering the rising poverty levels in Nigeria [46], there is a possibility that the public may opt for non-fluoridated toothpaste, posing a risk of increased caries prevalence, particularly among those who brush their teeth once daily. In response, the dental community should launch strategic campaigns to promote and encourage twice daily tooth brushing with fluoridated toothpaste. This proactive approach aims to mitigate the potential impact of the economic downturn on oral health.

Furthermore, the results of the current study suggest that the use of dental services by adolescents is poor. Currently, dental services are mainly sought for curative purposes, and adolescents lack awareness of preventive care benefits [47]. Barriers to healthcare service utilization in Nigeria include poor accessibility, cost, and a cultural reluctance towards preventive care [32, 48, 49]. A comprehensive, multifaceted response is essential to curb caries prevalence in a country like Nigeria where the oral health system is weak, poorly driving preventive care, and individual oral health behaviours of adolescents predisposes to high risk for caries.

Another area of concern is the data source utilized for caries prevalence studies in Nigeria, predominantly conducted in secondary schools. However, it is noteworthy that only 40% of eligible secondary school-age children are enrolled in school [49]. Moreover, adolescents aged 10 to 19 years exhibit diversity, spanning primary, secondary, and tertiary educational settings, with a considerable number already employed [50]. Furthermore, 44.0% of adolescent females are married, and 6.48% are mothers [51–53]. This local variability in the demographic profile of adolescents requires specific oral health interventions to address distinct needs. In addition, the social profile of female adolescents – early marriages [52–54] and a high rate of school dropouts [54] may present unique challenges affecting their oral health care and increasing susceptibility to caries. Consequently, the study underscores the importance of designing epidemiological studies that consider the local context. Such studies should explore effect of culture and context-based gender dynamics and the high prevalence of caries among females observed in this study, with the aim of designing gender-sensitive community-based oral health intervention programs for adolescents. This will help with the development of policies and programs tailored to the specific needs of the population.

In addition, for a country like Nigeria, population-based studies would be more appropriate for determining dental caries prevalence. Notably, only two out of the 16 studies included in this analysis were population-based. Furthermore, there was no implementation research exploring effective intervention models to respond to the oral health needs of adolescents; and caries prevalence data was concentrated in Southwest Nigeria. We also observed that data availability was biased towards regions in Nigeria hosting dental schools, with the data skewed to urban areas. These indicate the need for more comprehensive data on caries management for adolescents in Nigeria. Integrating oral health data into existing health surveillance systems, like the Demographic Health Survey [49], can provide a more inclusive assessment of caries profiles nationwide. Although the country has a national oral health policy [50], this is yet to translate to a dedicated national oral health program. Establishing and implementing a national oral health program can promote interventions aimed at improving the oral health of adolescents.

This pioneering study, on the caries profile among adolescents in Nigeria emphasizes an immediate requirement for a comprehensive public health strategy that addresses the high caries prevalence. Despite lower caries prevalence than the global average, there are concerns about the caries preventive behaviour that are suggestive of rising prevalence of dental caries in the absence of active interventions to eliminate the caries risk factors among adolescents. Although there is limited specific oral health data and programs for adolescents in Nigeria, prompt actions are needed towards limiting the potential negative impact which poor caries management may have on the mental, sexual, and reproductive health of adolescents. Multidimensional approaches are also crucial for culturally sensitive interventions and building responsive healthcare systems to manage dental caries among Nigerian adolescents.

## Conclusion

This systematic review and meta-analysis brings to light the high prevalence of dental caries that exist among Nigerian adolescents. It also emphasizes gender differences apparent in this area and the key preventive behaviours, like tooth brushing frequency. The study raises the issue of a need for a prompt and multidimensional public health response to this. High heterogeneity is a cautionary note about the variability and diversity within the study data and suggests a need for careful interpretation and consideration of the potential influencing factors.

## Data Availability

Not applicable

## Acknowledgement

Authors hereby acknowledge the Oral Health Initiative of the Nigeria Medical Research (NIMR) for providing grant for this study grant (Grant Number: OHI/COH2023/004).

